# Sex-Specific Clinical and Genetic Factors Associated with Adverse Outcomes in Hypertrophic Cardiomyopathy

**DOI:** 10.1101/2023.06.17.23291422

**Authors:** Alexandra Butters, Clare Arnott, Joanna Sweeting, Brian Claggett, Anna Cuomo, Dominic Abrams, Euan A. Ashley, Sharlene M. Day, Adam S. Helms, Rachel Lampert, Kim Lin, Michelle Michels, Erin M. Miller, Iacopo Olivotto, Anjali Owens, Victoria N. Parikh, Alexandre C. Pereira, Joseph W. Rossano, Thomas D. Ryan, Sara Saberi, John C. Stendahl, James S. Ware, John Atherton, Christopher Semsarian, Neal K. Lakdawala, Carolyn Y. Ho, Jodie Ingles

## Abstract

**Background:** Women with HCM present at a more advanced stage of the disease and have a higher risk of heart failure and death. The factors contributing to these differences are unclear. We aimed to investigate sex differences in clinical and genetic factors associated with adverse outcomes in the Sarcomeric Human Cardiomyopathy Registry (SHaRe).

**Methods:** Cox proportional hazard models were fit with a sex interaction term to determine if significant sex differences existed in the association between risk factors and outcomes. Models were fit separately for women and men to find the sex-specific hazard ratio.

**Results:** After a mean follow-up of 6.4 years, women had a higher risk of heart failure (HR 1.51; 95% CI 1.21-1.88, p=0.0003) but a lower risk of atrial fibrillation (0.74; 0.59-0.93; p<0.0001) and ventricular arrhythmia (0.60; 0.38-0.94; p=0.027) than men. No sex difference was observed for death (p=0.84). Sarcomere-positive men had a higher risk of heart failure and death, not seen in women (p-heterogeneity=0.006 & p-heterogeneity=0.035, respectively). *MYBPC3* variants were associated with lower heart failure risk in women than other HCM subgroups, with no significant change for men (p-heterogeneity <0.0001). Women with moderate risk of ventricular arrhythmia (4% to <6% ESC risk score) were at a higher risk of ventricular arrhythmia than those scoring <4%, not observed in men (p-heterogeneity= 0.019).

**Conclusion:** Clinical and genetic factors contributing to adverse outcomes in HCM affect women and men differently. Research to inform sex-specific management of HCM could improve outcomes for both sexes.

## INTRODUCTION

Hypertrophic cardiomyopathy (HCM) is the most common inherited heart disease affecting at least 1/500 in the general population.^1^ HCM is a heterogeneous condition, with approximately half of cases due to likely pathogenic or pathogenic (LP/P) autosomal dominant variants in genes encoding sarcomere proteins (sarcomere-positive), which are critical for contractile function of cardiomyocytes. The two most common genes implicated are myosin-binding protein C (*MYBPC3*) and β-myosin heavy chain (*MYH7*), which account for ∼75% of all HCM patients with causative variants ^2^. Despite the inheritance pattern conveying 50% risk to first-degree relatives, incomplete penetrance is common, and not all carriers of causative variants will develop disease.^3, 4^ There is increasing evidence that ∼40% have a non-Mendelian subtype, where the underlying disease mechanism is likely due to a combination of polygenic risk and comorbidities (sarcomere-negative).^5–8^

Published HCM cohorts are disproportionately male, with women accounting for only 30-45% of published study populations,^9^ despite this being considered an autosomal dominant disease. This might be explained by decreased disease penetrance in women, with male sex previously associated with three times the risk of developing HCM among individuals with sarcomere gene variants.^3^ Additionally, differences in clinical recognition due to the lack of sex-specific diagnostic criteria in clinical guidelines^17^ and unequal access to care also likely play a role.^10^ Indeed, multiple studies have also shown that women have a higher risk of adverse events, including heart failure and death.^10–13^ Despite the lower penetrance, women with HCM are more likely to have a LP/P sarcomere variant, while men are more likely to be sarcomere-negative.^5, 10^ Compared to men, women are older at diagnosis, present at an advanced stage of disease, have a higher prevalence of obstruction, and heart failure symptoms (New York Heart Association (NYHA) III-IV), worse exercise capacity and worse diastolic function.

We expand on previous work by evaluating sex-specific differences in risk factors for adverse outcomes in men and women with HCM. Early detection and management of cardiovascular risk factors remain vital for improving women’s cardiovascular health.^14^ We hypothesized that clinical and genetic factors contributing to adverse outcomes in HCM might be influenced differently by sex. Understanding the underlying factors contributing to these sex differences in outcomes of HCM is essential if we are to improve health outcomes and care for both men and women with HCM.

## METHODS

### Study Design

Details of SHaRe have been previously described.^7^ SHaRe is a consortium of patients from 12 international HCM referral centers. Each center maintains longitudinal databases capturing clinical, genetic and outcomes data on HCM patients and their families. Site data are mapped to corresponding fields in the secure, centralized database to produce standardized SHaRe definitions and values. Outcome events that occurred before or after the initial visit were identified and recorded. De-identified site data are mapped to corresponding discrete fields in the secure, centralized database to yield standardized SHaRe definitions and values. Prospective data are captured via quarterly uploads from site databases. Each participating site has received ethical approval in accordance with local policies, as follows: Ethical approval requiring participant informed consent was granted by Cincinnati Children’s Hospital, United States of America (USA); Children’s Hospital of Philadelphia, USA; Michigan Medical, USA; Yale Medical, USA; Royal Brompton Hospital, United Kingdom; Erasmus University Medical Center, The Netherlands; Florence Centre for Cardiomyopathies, Italy; and Sydney Local Health District, Royal Prince Alfred Hospital, Australia; InCor, Heart Institute, University of Sao Paulo, Brazil institutional review boards. Ethical approval with a waiver of consent was granted by Stanford School of Medicine, USA; Brigham and Women’s Hospital, USA; Boston Children’s Hospital, USA; Pennsylvania University Medical Center, USA; and Sydney Local Health District, Royal Prince Alfred Hospital, Australia institutional review boards. All individual-level data are pseudonymized prior to upload to SHaRe.

### Study population

Patients were included if they had a site-designated diagnosis of HCM, defined as unexplained left ventricular (LV) hypertrophy with maximal wall thickness >13mm if positive genetic testing, or >15mm if negative genetic testing and negative family history, or equivalent z score for pediatric patients. Only probands were included, i.e., the first affected family member to seek medical attention for HCM. All analyzed metrics of cardiac dimensions and function were based on echocardiographic measurements.

### Genetic testing

Genetic testing was performed at all sites using different tests available over time. Variants were classified as pathogenic (P), likely pathogenic (LP), unknown significance (VUS), or benign/likely benign by each site using contemporary criteria, concentrating on the eight sarcomere genes definitively associated with HCM (myosin binding protein C [*MYBPC3*], myosin heavy chain [*MYH7*], cardiac troponin T [*TNNT2*], cardiac troponin I [*TNNI3*], α-tropomyosin [*TPM1*], myosin essential and regulatory light chains [*MLY2, MYL3*], and actin [*ACTC1*]). [3]. Sarcomere variants with ambiguous classifications underwent additional systematic review by a subgroup of investigators to adjudicate and standardize classification (SD, JW, JI). Patients were excluded if they had potentially pathogenic variants in genocopy genes such as α-galactosidase (*GLA*) or lysosome-associated membrane protein (*LAMP2*), indicating the presence of metabolic or storage disease or diagnoses other than HCM. Genotyped patients were designated as sarcomere-positive (at least one LP/P variant in any of the above sarcomere genes), sarcomere-negative (no causative sarcomere variants identified), or VUS (variant of uncertain significance in a sarcomere gene).

### Outcome definitions

The four primary adverse outcomes were heart failure composite, ventricular arrhythmia composite, atrial fibrillation (AF) and death. Outcomes were documented by site investigators during routine clinical encounters and captured directly into the database. Composite outcomes were defined as follows:

*Heart failure composite:* the first occurrence of cardiac transplantation, LV assist device implantation, or NYHA functional class III-IV symptoms.

*Ventricular arrhythmia composite:* the first occurrence of sudden cardiac death, resuscitated cardiac arrest, or appropriate ICD therapy.

### Statistical analysis

Baseline characteristics for women and men are presented as number (percentage) for categorical variables, mean (standard deviation) for approximately symmetrical continuous variables, and median (interquartile range) for asymmetrical continuous variables. Continuous variables were compared using the unpaired t-test, and categorical variables using the χ2 test. The incidence of adverse outcomes (heart failure composite, ventricular arrhythmia composite, AF, death) was estimated separately for women and men. Multivariable Cox proportional hazard models were used to estimate hazard ratios (HR) and 95% confidence intervals (CI) for adverse outcomes comparing women with men. Only incident outcomes were included in analyses and the time variable was from first evaluation at a SHaRe site to either development of the outcome, or last follow-up for those who had no events. All models were adjusted by age at the first encounter at a SHaRe center. Patients missing data on the occurrence or timing of events were not included in analyses of those outcomes. However, to maximize sample sizes across analyses, such patients were included in analyses of other outcomes where data were available.

The association between baseline risk factors and adverse outcomes was assessed using Cox regression models. Heterogeneity of associations was tested between women and men by adding an interaction term into the Cox regression model with no adjustment for multiplicity. Time was from first evaluation at a SHaRe site to either development of the outcome, or last follow-up for those who had no events. All models were adjusted by age at the first encounter at a SHaRe site, and echocardiographic models were adjusted for body surface area. Data were reported as HR with 95% CI for women versus men as the reference. A 2-sided p-value <0.05 was considered statistically significant. Multiple corrections were not applied as this is hypothesis generating research. Kaplan[Meier survival analysis with log-rank comparison was used to compare the age of heart failure composite and death separately for women and men by sarcomere status and genotype. The time variable was age (years) at first event or age at most recent follow-up where they were known to be free from events, ≤80 years. Longitudinal analysis of echocardiographic parameters was performed using linear mixed models (Supplementary material). All analyses were conducted using R version 4.1.2.

## RESULTS

### Demographic and clinical characteristics

There were 7315 patients with HCM seen between 1960-2020, and after excluding 768 family members, there were 6647 probands included in the study (Figure 1A). There were 2538 (38%) women, with an overall mean follow-up time from the first to the last encounter of 6.4 ± 7.1 years (Table 1). The cohort consisted of 667 pediatric patients (<18 years), of which 226 (33.9%) were female and 441 (66.1%) male.

**Figure 1:**
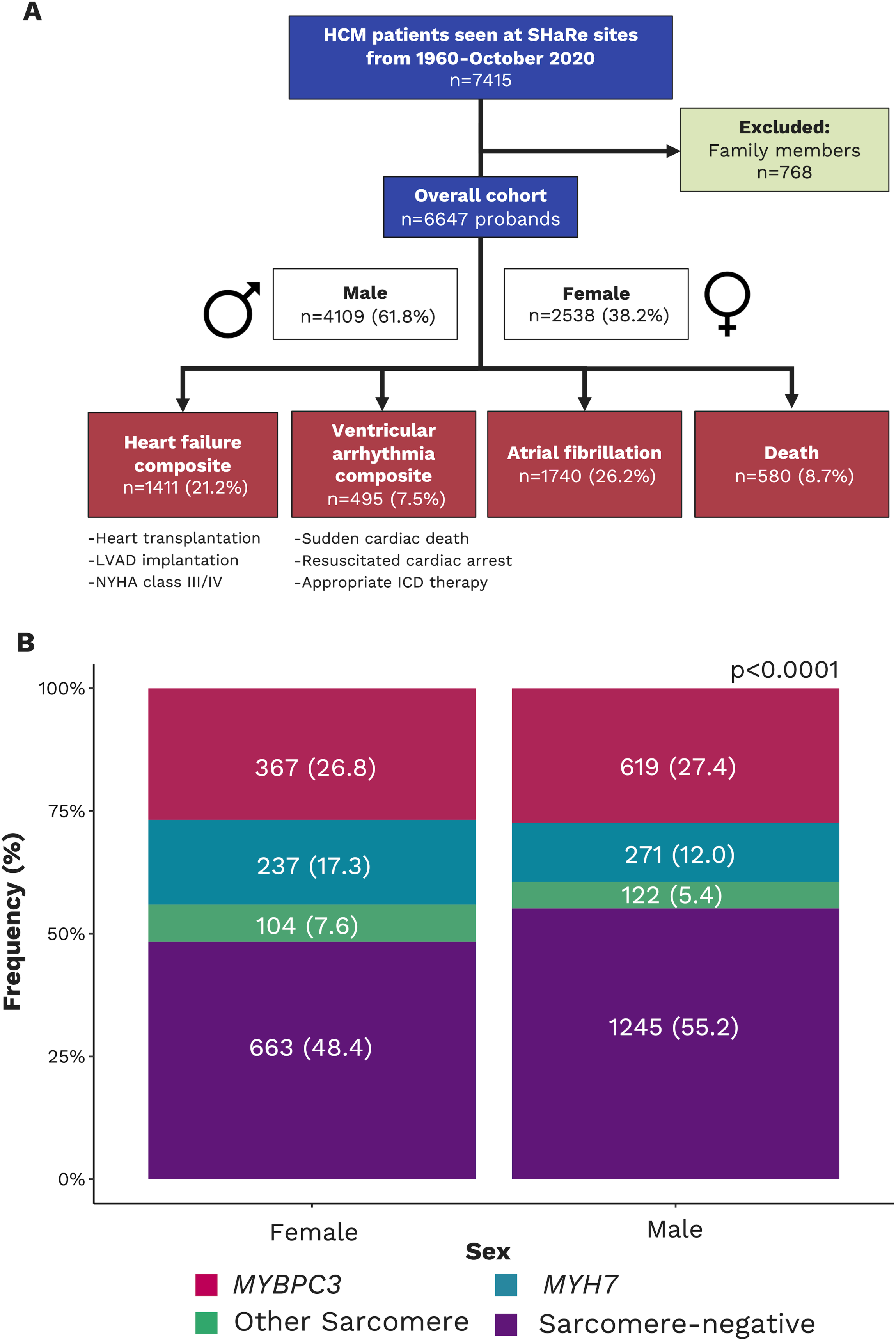
Study outcomes and genetic findings. **(A)** Flow diagram of study participants **(B)** Sarcomere variant gene distribution including sarcomere-negative probands. Abbreviations: ICD, implantable cardioverter defibrillator; LVAD, left ventricular assist device; NYHA, New York Heart Association.

**TABLE 1:** Demographic and baseline characteristics of hypertrophic cardiomyopathy probands by sex.

|  | Total cohort | Female | Male | p-value |
| --- | --- | --- | --- | --- |
|  | 6647 | 2538 (38.2) | 4109 (61.8) |  |
| Age at diagnosis (years) | 43.9 ± 19.5 | 47.5 ± 20.6 | 41.6 ± 18.4 | <0.0001 |
| Follow-up time (years) | 6.4 ± 7.1 | 6.3 ± 6.8 | 6.5 ± 7.2 | 0.412 |
| Race |  |  |  |  |
| White | 5465 (91.8) | 2114 (92.2) | 3351 (91.6) | 0.300 |
| Black | 269 (4.5) | 92 (4.0) | 177 (4.8) |  |
| Asian | 217 (3.7) | 87 (3.8) | 130 (3.6) |  |
| Genetic testing | 4182 (62.9) | 1552 (61.2) | 2630 (64.0) | 0.019 |
| Sarcomere status |  |  |  | <0.0001 |
| Positive | 1775 (48.2) | 728 (52.3) | 1047 (45.7) |  |
| Negative | 1908 (51.8) | 663 (47.7) | 1245 (54.3) |  |
| Gene status |  |  |  | 0.001 |
| *MYBPC3* | 986 (57.3) | 367 (51.8) | 619 (61.2) |  |
| *MYH7* | 508 (29.5) | 237 (33.5) | 271 (26.8) |  |
| Other sarcomere | 226 (13.1) | 104 (14.7) | 122 (12.1) |  |
| Hypertension | 2690 (40.5) | 1091 (43.0) | 1599 (38.9) | 0.001 |
| Body mass index (kg/m<sup>2</sup>) | 27.5 ± 6.2 | 27.3 ± 7.0 | 27.7 ± 5.7 | 0.080 |
| NYHA III-IV | 792 (15.3) | 455 (22.9) | 337 (10.5) | <0.0001 |
| Syncope | 1008 (15.6) | 432 (17.5) | 576 (14.4) | 0.001 |
| **Echo characteristics** |  |  |  |  |
| Indexed Max LV wall thickness, mm<sup>a</sup> | 9.9 ± 2.3 | 10.4 ± 4.3 | 9.5 ± 4.2 | <0.0001 |
| Indexed LV end diastolic diameter, mm<sup>a</sup> | 23.6 ± 6.6 | 24.2 ± 6.7 | 23.2 ± 6.6 | <0.0001 |
| Indexed end systolic diameter, mm<sup>a</sup> | 14.3 ± 5.2 | 14.5 ± 2.9 | 14.2 ± 5.4 | 0.071 |
| LV ejection fraction (%) | 64.9 ± 9.3 | 65.6 ± 9.8 | 64.5 ± 9.0 | <0.0001 |
| LV outflow tract obstruction | 1981 (40.2) | 895 (46.9) | 1086 (36.0) | <0.0001 |
| LV outflow tract gradient at rest (mmHg) | 30.8 ± 34.6 | 37.1 ± 38.4 | 26.9 ± 31.4 | <0.0001 |
| Mitral regurgitation | 3737 (64.6) | 1527 (68.6) | 2210 (62.2) | <0.0001 |
| Indexed left atrial diameter, mm<sup>a</sup> | 22.6 ± 6.2 | 23.9 ± 6.5 | 21.7 ± 5.8 | <0.0001 |
| **ESC HCM Risk score** |  |  |  |  |
| Low risk (<4%) | 4220 (87.0) | 1627 (89.2) | 293 (85.7) | 0.0002 |
| Moderate risk (4 to <6%) | 415 (8.6) | 117 (6.4) | 298 (9.9) |  |
| High risk (≥6%) | 216 (4.5) | 81 (4.4) | 135 (4.5) |  |

Data shown as n (%) or mean ± SD. ESC, European Society of Cardiology; NYHA indicates New York Heart Association; LV, left ventricular
<sup>a</sup>Adjusted for body surface area

**TABLE 2:** Incident medication use, interventions and outcomes by sex.

|  | **Total cohort** | **Women** | **Men** |  |
| --- | --- | --- | --- | --- |
| **Medications** | 6647 | 2538 (38.2) | 4109 (61.8) | p-value |
| Beta blocker | 3789 (60.1) | 1495 (62.2) | 2294 (58.8) | 0.001 |
| Anti-arrhythmic | 718 (13.7) | 275 (13.8) | 443 (13.7) | 0.953 |
| Calcium channel blocker | 1211 (19.3) | 529 (22.0) | 682 (17.6) | <0.0001 |
| Disopyramide | 476 (8.4) | 240 (11.0) | 236 (6.7) | <0.0001 |
| Loop diuretic | 308 (10.4) | 157 (14.7) | 151 (7.9) | <0.0001 |
| **Interventions** |  |  |  |  |
| ICD implantation | 1697 (25.5) | 633 (24.9) | 1064 (25.9) | 0.386 |
| Alcohol septal ablation | 295 (4.5) | 160 (6.3) | 135 (3.3) | <0.0001 |
| Septal myectomy | 1124 (16.9) | 505 (19.9) | 619 (15.1) | <0.0001 |
| **Outcomes** |  |  |  |  |
| Heart failure composite | 1411 (21.2) | 729 (28.7) | 682 (16.6) |  |
| Ventricular arrhythmia composite | 495 (7.5) | 175 (6.9) | 320 (7.8) |  |
| Atrial fibrillation | 1740 (26.2) | 681 (26.8) | 1059 (25.8) |  |
| Death | 580 (8.7) | 289 (11.4) | 291 (7.1) |  |

Data shown as n (%) or mean ± SD. ICD indicates implantable cardioverter defibrillator.
Heart failure composite: first occurrence of cardiac transplantation, left ventricular assist device implantation, or New York Heart Association functional class III-IV symptoms.
Ventricular arrhythmic composite: first occurrence of sudden cardiac death, resuscitated cardiac arrest, or appropriate ICD therapy.

Women were approximately 6.0 years older than men at diagnosis (47.5 ± 20.6 versus 41.6 ± 18.4; p<0.0001). At baseline, NYHA class III-IV symptoms were present in 455 (22.9%) of women, compared to only 337 (10.5%) of men; p<0.0001. Additionally, 895 (47%) women had LV outflow tract obstruction (LVOTO) (defined as LVOTO ≥30mmHg at rest or with provocation) compared to 1086 men (36%); p<0.0001). Subsequently, women also had significantly higher LV outflow tract gradient 37.1 ± 38.4 versus 26.9 ± 31.4; p<0.0001). LV ejection fraction was greater in women, although not clinically significant (65.6 ± 9.8 versus 64.4 ± 9.0; p<0.0001). After adjusting for body surface area, women had significantly greater absolute maximum LV wall thickness, LV cavity diameter, and left atrial (LA) diameter (Table 1).

Genetic testing was performed in 4182 (63%) probands, with a total of 2630 (64%) men and 1552 (61%) women undergoing genetic testing (p=0.019) (Table 1). Sarcomere status differed by sex; a total of 728 (52%) women and 1047 (46%) men were sarcomere-positive (p<0.0001). Among the sarcomere-positive group, most had *MYBPC3* variants, although men were proportionately even more likely to have *MYBPC3* variants (p=0.001). Proportionately more women had variants in *MYH7* and other sarcomere genes compared to men (p=0.001; Figure 1B).

### Incident use of medications and interventions

Table 1 shows overall medication use and interventions in the cohort. Compared to men, women were more likely to be prescribed beta-blockers (1495 [63%] women versus 2294 [59%]; p=0.001), calcium channel blockers (529 [22%] versus 682 [18%]; p<0.0001), disopyramide (240 [11%] versus 236 [7%]; p<0.0001), and loop diuretics (157 [15%] versus 151 [8%]; p<0.0001), though noting data missingness for these variables (missing, 5%-14%), especially loop diuretics (55%). There was no sex difference in the prescription of anti-arrhythmic drugs (p=0.953). The prevalence of ICD insertion was similar between the sexes (p=0.386). In patients with obstruction, women were significantly more likely than men to undergo alcohol septal ablation (102/895 [11.4%] versus 80/1082 [7.4%] p=0.002); however, there was no sex difference for septal myectomy (331/895 [37.0%] versus 361/1086 [33.2%]; p=0.08).

### Adverse clinical outcomes

During the follow-up period, a total of 1411 heart failure composite events were recorded (729, 51.7% women), 495 ventricular arrhythmia composite events (175, 35.4% women), 1740 AF events (1740, 39.1% women), and 580 deaths (289, 49.8% women; Table 1). Multivariate Cox models were fit to test if sex is an independent predictor of key outcomes (Supplemental Table 1). Women had a higher risk of heart failure (HR 1.51, CI 1.21-1.88; p=0.0003) in a model adjusted for age, sarcomere status, baseline LVEF<50%, baseline indexed LA size, baseline LVOTO, body mass index (BMI) and hypertension. However, female sex did not remain a significant predictor of death in a model adjusted for age, sarcomere status, baseline LVEF<50%, baseline indexed LA size and baseline LVOTO (HR 0.96, CI 0.66-1.41; p= 0.843). Additionally, women had a reduced risk of AF (HR 0.74 CI 0.59-0.93; p<0.0001) and ventricular arrhythmia (HR 0.60 CI 0.38-0.94; p=0.027) compared to men.

### Sex-specific clinical and genetic factors for heart failure composite

Sex differences existed for the associations between the heart failure composite and sarcomere-positive status, LP/P variants in *MYBPC3*, and baseline indexed LA diameter after adjusting for age at first encounter (Figure 2). In men, sarcomere-positive status was associated with an increased risk of the heart failure composite compared to sarcomere-negative patients (HR 1.34 [95% CI 1.06-1.69]), while for women, there was no significant change in risk by sarcomere status (HR 0.85 [95% CI 0.66-1.08]; p-heterogeneity=0.006). However, having an LP/P variant in *MYBPC3* was associated with a lower risk of the heart failure composite for women compared to sarcomere-negative HCM (HR 0.56 [95% CI 0.41-0.77]), with no significant change for men (HR 1.29 [95%CI 0.99-1.67]; p-heterogeneity <0.0001).

**Figure 2:**
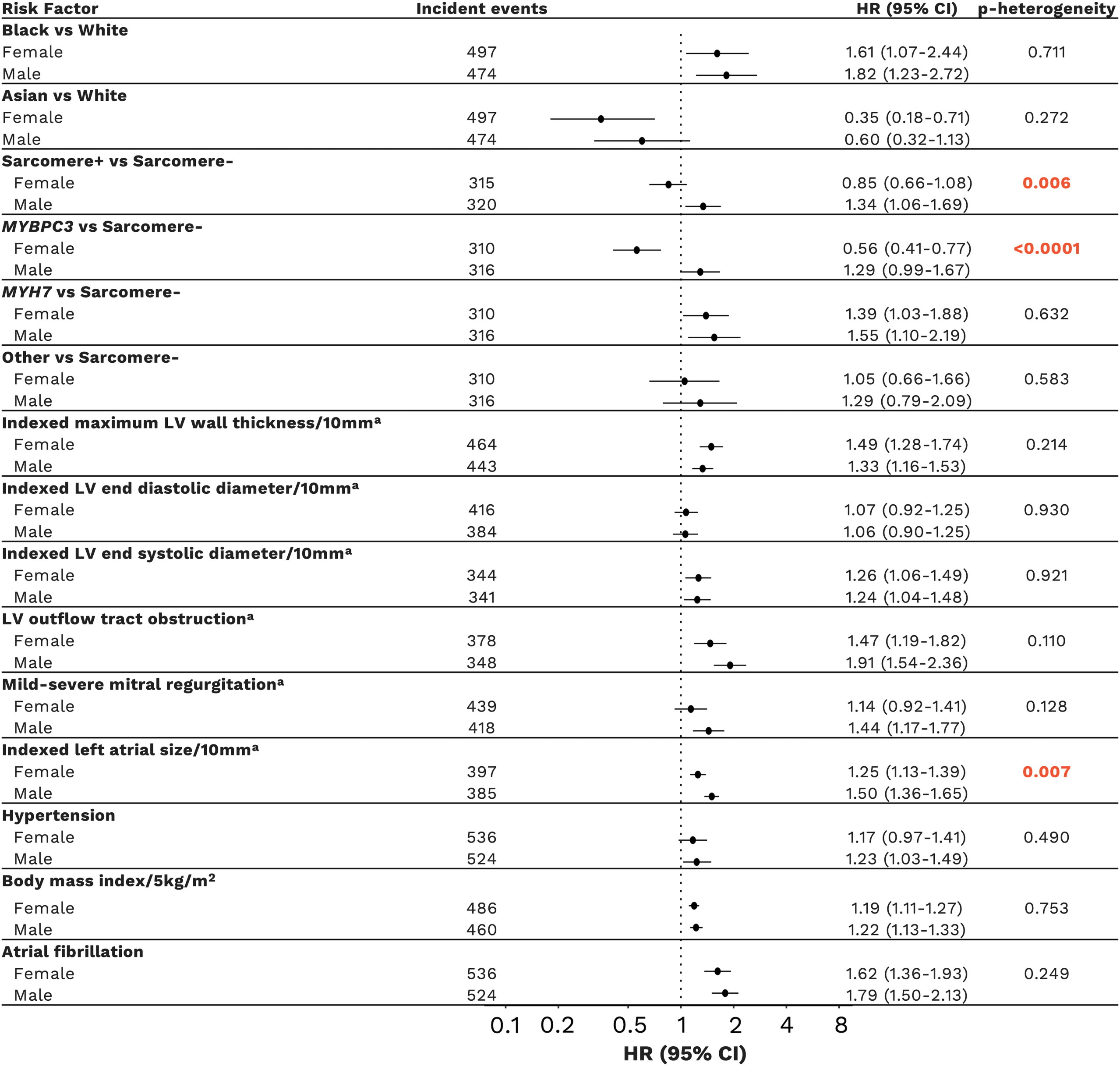
Sex-disaggregated analysis showing clinical and genetic features for heart failure. All models were adjusted for age at first evaluation. ^a^Echocardiogram measurements adjusted for body surface area. Abbreviations: CI; confidence interval; HR; hazard ratio; LV, left ventricular.

LV ejection fraction <35% at baseline was associated with an increased risk of the heart failure composite in both women and men; however, men had a much higher relative risk (HR 12.64 [95% CI 7.29-21.92]) compared to women (HR 3.46 [95% CI 1.82-6.55]; p-heterogeneity 0.002). Greater indexed LA diameter (per cm increase) was associated with an increased risk of the heart failure composite in both women and men, but slightly more so in men (HR 1.50 [95% CI 1.36-1.65] compared to women (HR 1.25 [1.13-1.39]; p-heterogeneity 0.007).

While black race, LP/P variants in *MYH7*, larger baseline indexed maximum LV wall thickness, larger baseline indexed LV end-systolic diameter, baseline LV outflow tract obstruction, LV ejection fraction <50%, greater BMI, and AF all increased the risk of the heart failure composite in the cohort, there was no heterogeneity by sex for any of these associations (p-heterogeneity >0.05).

### Sex-specific clinical and genetic factors for death

Sex differences existed for the associations between death and sarcomere-positive status, greater BMI, and the heart failure composite after adjusting for age at first encounter (Figure 3). Compared to sarcomere-negative HCM, sarcomere-positive status increased the risk of death in men (HR 1.48 [95% CI 1.08-2.04]) but not in women (HR 0.86 [95% CI 0.71-1.21]; p-heterogeneity=0.035). Greater BMI increased the risk of death in women (HR 1.18 [95% CI 1.05-1.31]) but not in men (HR 0.96 [95% CI 0.82-1.12]; p-heterogeneity 0.031). Heart failure increased the risk of death to a greater extent in men (HR 2.18 [95% CI 1.72-2.78]) than in women (HR 1.45 [95% CI 1.14-1.83]; p-heterogeneity=0.013).

**Figure 3:**
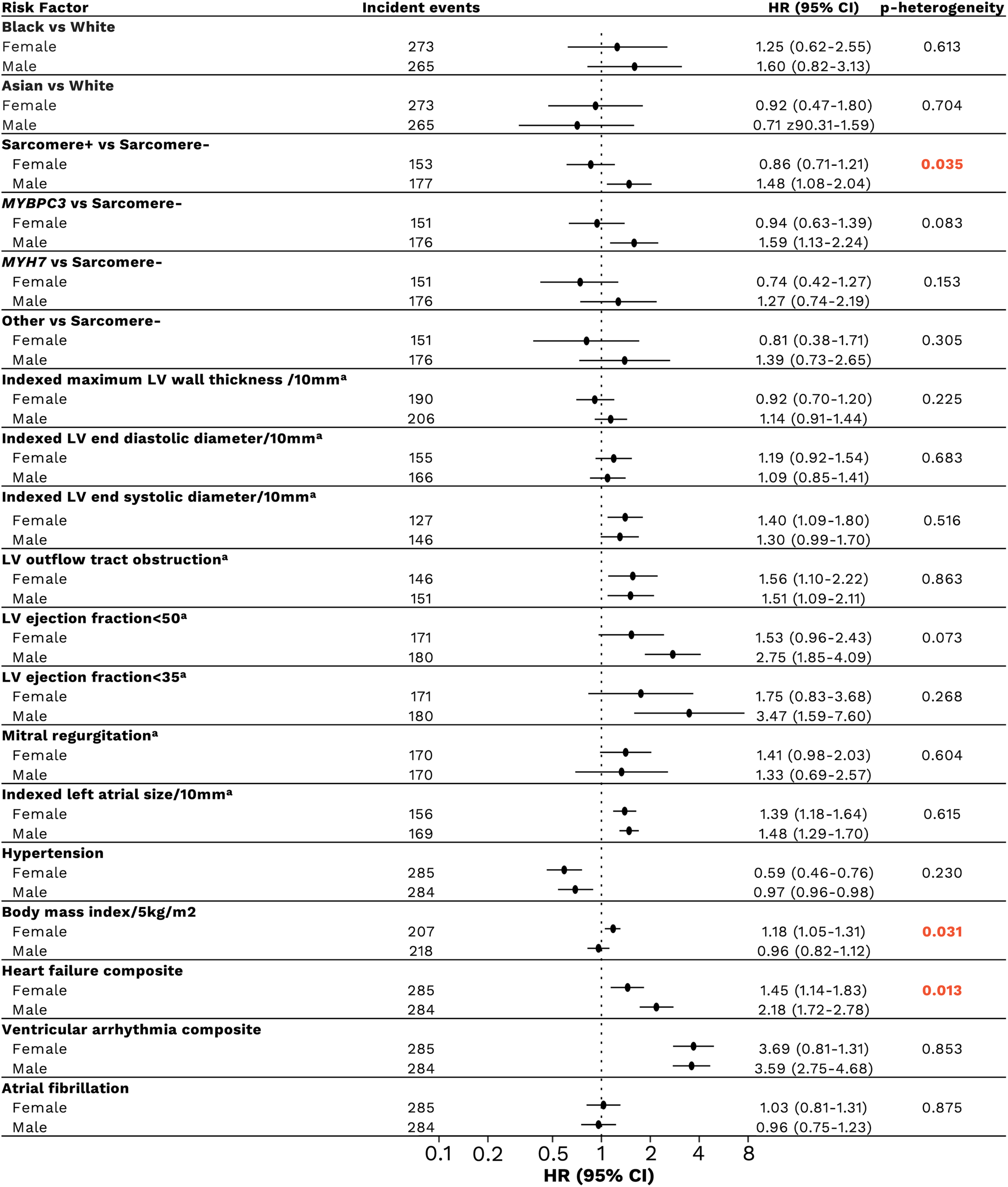
Sex-disaggregated analysis showing clinical and genetic features for death. All models were adjusted for age at first evaluation. ^a^Echocardiogram measurements adjusted for body surface area. Abbreviations: CI; confidence interval; HR; hazard ratio; LV, left ventricular.

The risk of death was increased for both sexes for the associations between baseline LV outflow tract obstruction, baseline indexed LA diameter, and ventricular arrhythmia composite. Hypertension was consistently associated with a lowered risk of death in both sexes (all p-heterogeneity >0.05).

### Sex-specific clinical and genetic factors for AF

No heterogeneity by sex was observed in associations between risk factors and AF after adjusting for age at first encounter (Supplemental Figure 1). Sarcomere-positive status, disease-causing variants in *MYH7*, larger baseline indexed maximum LV wall thickness, baseline LV outflow tract obstruction, mitral regurgitation, baseline indexed LA diameter and greater BMI were all associated with an increased risk of AF in both sexes (p-heterogeneity >0.05).

### Sex-specific clinical and genetic factors for ventricular arrhythmia composite

Sex differences were identified for the associations between the ventricular arrhythmia composite and European Society of Cardiology (ESC) HCM risk score and greater baseline BMI after adjusting for age at first encounter (Figure 4). There was a sex difference in the association between moderate ESC HCM risk (4 to <6%) compared to low risk (<4%), with women at moderate risk more likely to experience ventricular arrhythmia (HR 3.57 [95% CI 1.70-7.49]), and this was not observed in men (HR 1.13 [95% CI 0.63-2.03]; p-heterogeneity=0.019). Men had a 1.3-fold increased risk of ventricular arrhythmia composite for every 5kg/m^2^ increase in BMI above the mean (HR 1.29 [95% CI 1.14-1.48]) with no significant change for women (HR 1.04 [95% CI 0.87-1.24]; p-heterogeneity 0.030).

**Figure 4:**
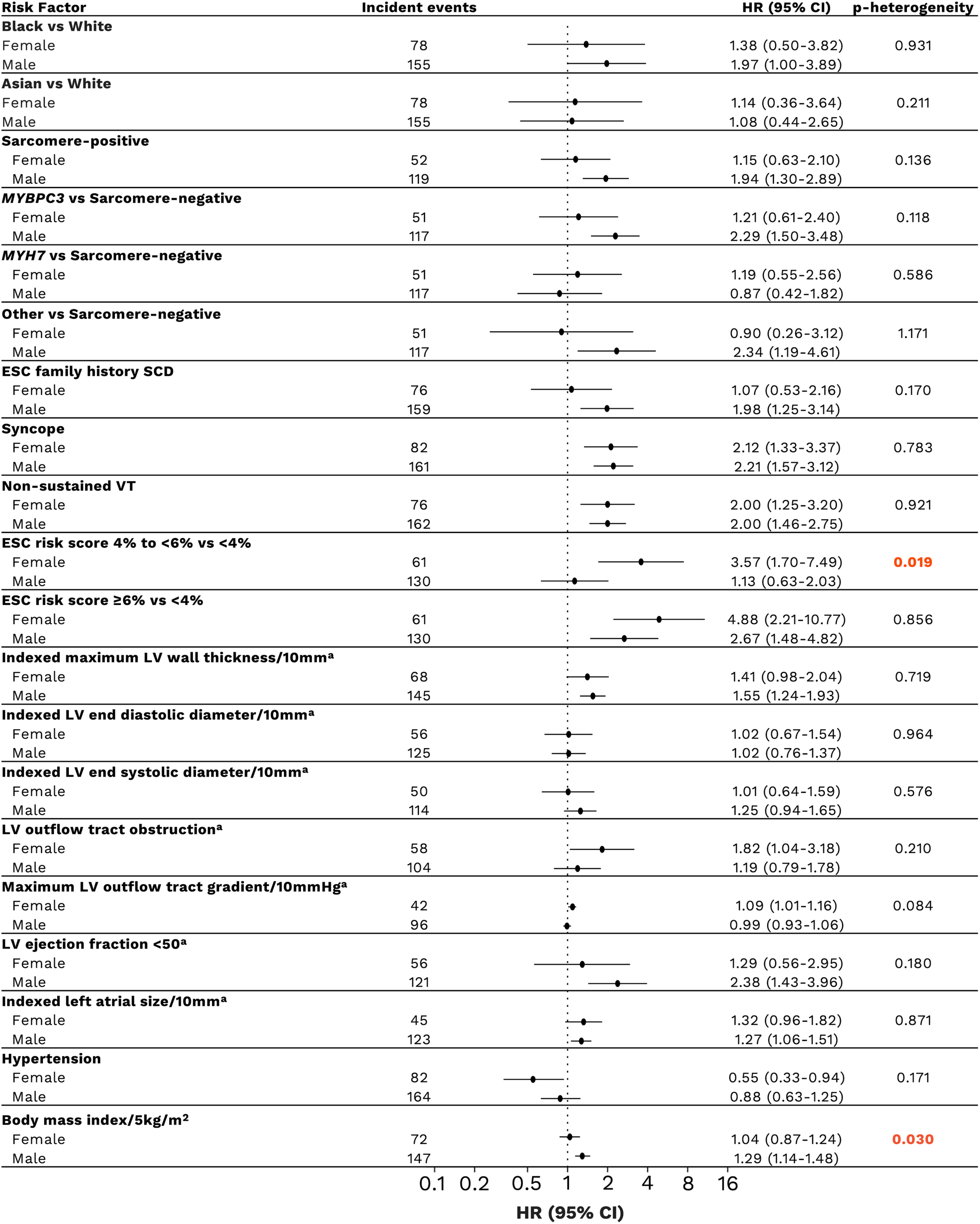
Sex-disaggregated analysis showing associations of clinical and genetic features with ventricular arrhythmia. ^a^Echo measurements adjusted for body surface area Abbreviations: HR; hazard ratio, CI; confidence interval; ESC, European Society of Cardiology; SCD, sudden cardiac death; LV, left ventricular.

The risk of ventricular arrhythmia was increased for both sexes for the associations between high ESC HCM risk score (>6%), non-sustained ventricular tachycardia and syncopal events, with no heterogeneity by sex (p-heterogeneity >0.05).

### Increased risk of events over time for sarcomere-negative women

To further explore why genotype is a sex-specific risk factor, Kaplan[Meier survival analysis with log-rank comparison was used to evaluate time to event for each sex descriptively. While sarcomere-negative men and women were initially less likely to develop heart failure compared with sarcomere-positive individuals, this difference diminished over time in women but persisted in men (Figure 5a). However, the risk remained higher in men and women with *MYH7* variants and men with *MYBPC3* variants (Figure 5b). Regarding death, for both sexes, those who are sarcomere-positive, on average, die at a younger age than sarcomere-negative individuals (Figure 5c). A comparison of the baseline characteristics, medications and interventions between sarcomere-positive and sarcomere-negative patients by sex can be found in Supplementary Table 2 & 3.

**Figure 5:**
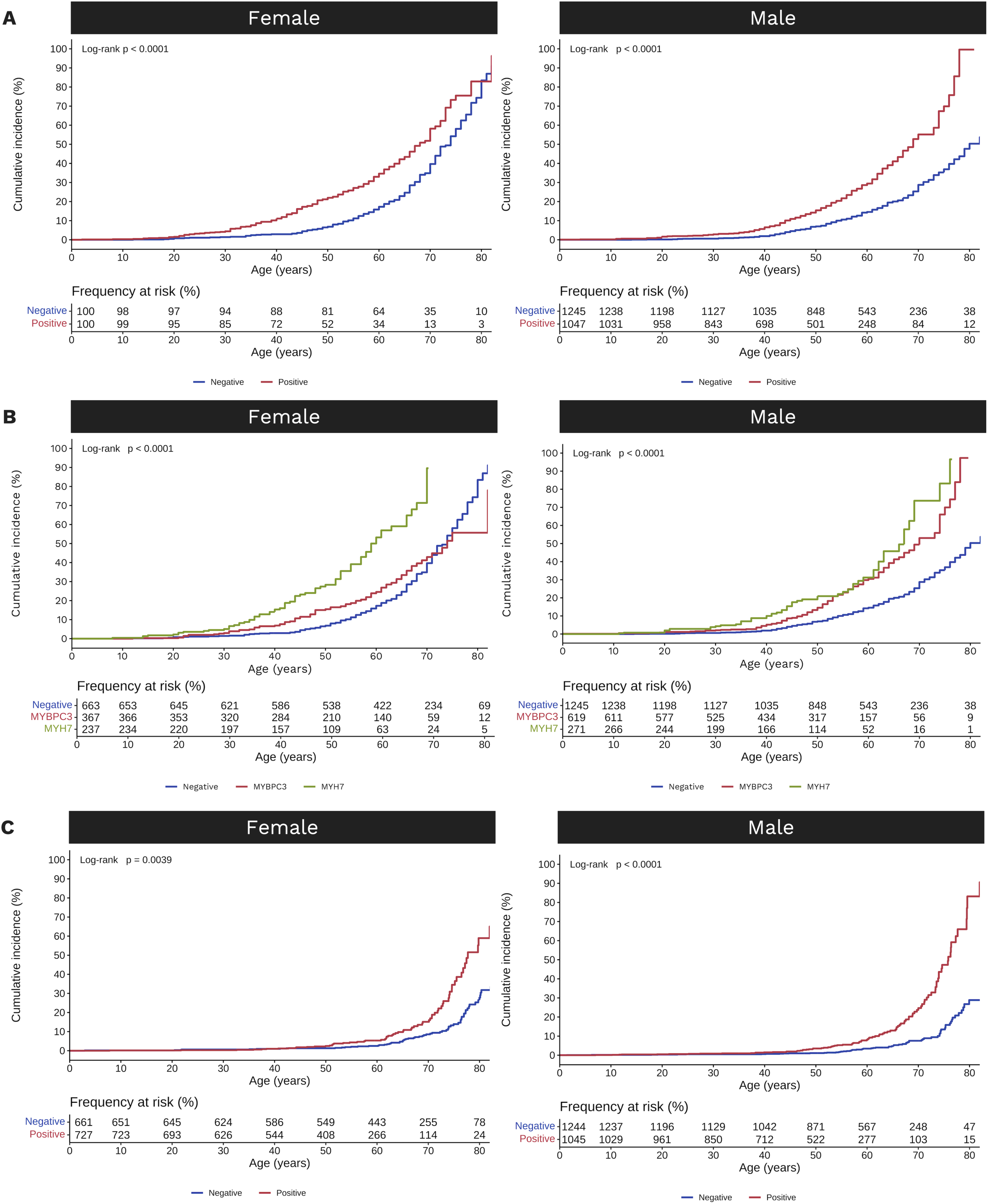
Kaplan□Meier survival analysis showing the difference in lifetime risk of **(A)** the heart failure composite for males and females by sarcomere status; **(B)** the heart failure composite for males and females by gene status; and **(C)** death by sarcomere status for females and males.

## DISCUSSION

The majority of clinical evidence informing the management of HCM is predominantly based on studies involving men.^9, 15^ It is therefore essential to gain a more comprehensive understanding of the social, clinical and biological factors that differentiate between sexes. A critical first step in addressing sex disparities is the reporting of sex-disaggregated statistics.^16^ We show important sex differences in risk factors associated with heart failure, death and ventricular arrythmia in HCM, but not AF (Figure 6). Notably, we observed that although sarcomere-negative men and women were initially less susceptible to developing heart failure compared with sarcomere-positive individuals, sarcomere-negative women had an increased risk of heart failure later in life, similar to the heightened incidence observed in the general female population. Additionally, the presence of *MYBPC3* variants in women decreased the risk of heart failure in younger women, compared to men with *MYBPC3* variants, suggesting sex-based mechanistic differences. Importantly, the impact of AF, BMI, LV wall thickness and obstruction on heart failure, and to large extent, death, were by similar sex. We show sex differences in ESC HCM risk score in predicting risk of ventricular arrhythmia, whereby women with moderate ESC HCM risk had an increased risk of ventricular arrhythmia compared to women at low ESC HCM risk. No significant difference in risk was observed for men. As the risk score does not consider sex, this may indicate risk for women is not adequately predicted using this tool. Our findings underscore the need for pre-specified sex-disaggregated analyses to elucidate sex differences and suggest the potential value of sex-specific medical management recommendations that may improve outcomes. Further research, validation of our findings, and a better understanding of the role of genetics in predicting sex-based risk will inform a precision-based approach to care that benefits both men and women.

**Figure 6:**
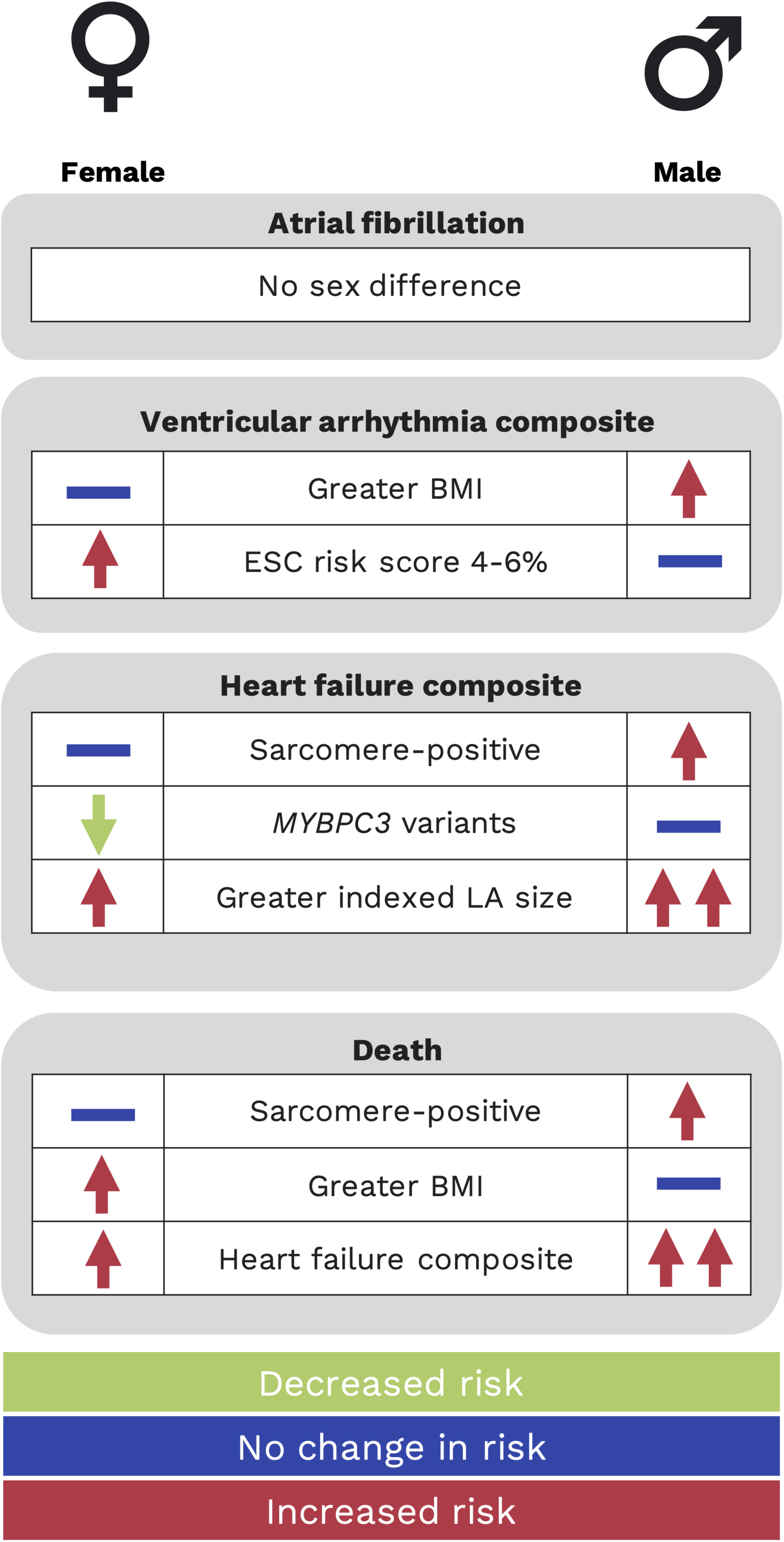
Sex-specific risk factors for adverse outcomes in hypertrophic cardiomyopathy. Abbreviations: BMI, body mass index; EF, ejection fraction; LA, left atrial.

The underlying genetic basis of HCM has a differential impact on adverse outcomes across the sexes. We demonstrated that LP/P variants in sarcomere genes increased the risk of heart failure and death for men but not women. Unlike sarcomere-negative men, sarcomere-negative women had an increased risk of heart failure and death in later life, likely mirroring the increasing risk of heart failure in women seen in the general population in later life.^17^ In fact, the prevalence of heart failure increases with age for both sexes in the general population, but by the age of 79 more women than men have heart failure, with more associated comorbidities but better survival than men.^18–20^ Sarcomere-negative men had a lower lifetime risk of both heart failure and death compared to sarcomere-positive men, consistent with previous reports in the general HCM literature, suggesting a more benign clinical course compared to sarcomere-positive men with HCM.^5–7^ Recent work to understand the underlying disease etiology in this group has shown an important polygenic basis, and diastolic blood pressure has been demonstrated as an independent contributor to disease development in sarcomere-negative HCM.^21, 22^ Interestingly, important sex differences exist for blood pressure in the general population, with men having a greater prevalence of hypertension overall, but women experiencing steeper increases in measures earlier in life, hypothesized to contribute to cardiovascular disease risk in later life for women.^23, 24^ The observed sex differences among sarcomere-negative patients in our study and known differences in modifiable risk factors such as hypertension signal a critical need for future studies to report sex-disaggregated data and consider clinical implications for each sex.

LP/P variants in *MYH7* and *MYBPC3* account for the majority of sarcomere-positive HCM, and we highlight important sex differences at the gene level. Men and women with *MYH7* variants had a greater risk of heart failure than those with sarcomere-negative HCM.

However, among women, LP/P variants in *MYBPC3* did not similarly increase risk. Although, subtle genotype-phenotype associations exist in HCM, such as LP/P variants in *MYH7* associated with a younger age at presentation^25^ and greater risk of AF,^26, 27^ the overall marked phenotypic and genetic heterogeneity in HCM limits the clinical utility of this information. Variants in *MYBPC3* are frequently truncations, with a proportion being founder variants.^28^ Founder variants, which must withstand negative selection pressure to be inherited over generations, were initially considered to cause less penetrant or milder disease. However, more recent data suggest they do not confer a more benign prognosis ^28, 29^, with recent SHaRe data showing founder truncating variants have an identical effect as non-founder truncating variants ^29^. Therefore, the significance of this finding in our study is unclear. Nevertheless, it raises interesting questions about the impact of the underlying biological cause of disease and its potential modification by sex.

Other adverse outcomes, including ventricular arrhythmia, AF and death, were also evaluated in this study. We showed a sex-specific difference in risk of ventricular arrhythmia between women and men with a moderate ESC HCM Risk score, compared to those with a low risk. Considering the key variables comprising the score, which do not include sex, this finding suggests the risk calculator might not adequately discriminate between those at high and low risk of sudden cardiac death in women. Men with greater BMI had an increased risk of the ventricular arrhythmia composite, while no difference was seen for women. Studies investigating sudden cardiac death in the general population have likewise reported this association previously, although there is disagreement on whether this is due to increased myocardial hypertrophy and fibrosis, which could potentially be a mechanism for ventricular arrhythmia.^30, 31^ Comorbidities and other modifiable risk factors significantly contribute to cardiovascular disease risk in both men and women in the general population.^14^ Although these data were limited in our HCM dataset, a more extensive investigation of both traditional, sex-specific and non-traditional risk factors in HCM could provide even greater clarity on sex differences. Given the potential role of sex-specific hormones in modulating disease, gathering female-specific data, such as menarche, pregnancy and reproductive-associated variables, could be particularly important.

While the development of heart failure is a true finding, the fact that more women have worse heart failure at initial visit, may be partly due to referral bias. Women are often sicker at presentation because they are not referred for evaluation, particularly to specialty centers, as early as men.^32–34^ Women typically have smaller hearts than men and consequently, women require a more significant degree of hypertrophy to fulfil the diagnostic criteria of maximal wall thickness of at least 15mm (or 13mm in first-degree relatives). This underlying difference in biology has been postulated to contribute to the delay in diagnosis and the higher initial diagnosis of peak LV outflow gradient observed in women.^35^ A general assumption that men are more at risk of cardiovascular disease, leading to reduced awareness and prioritization among women must also be considered. The Heart Foundation of Australia reported that only 39% of women aged ≥45 years had undergone a heart risk assessment by a healthcare professional in the previous two years.^36^ Indeed, physician bias contributes to reduced screening and prevention efforts for women compared to men.^37^ Our results show equal access and utilization of medical therapy and interventions by sex when treated in a SHaRe center. Investigation of why women are referred to specialist centers later than men is warranted.

## LIMITATIONS

Use of the heart failure composite outcome provides an incomplete measure of heart failure burden in HCM, including NYHA class III-IV symptoms which could be attributed to other causes such as LV obstruction. We had incomplete and limited information about comorbidities and could not interrogate the impact of menarche, pregnancy or reproductive history. Further research is necessary to determine the effects of these factors on adverse outcomes in HCM. Finally, all patients come from specialized HCM clinics that have been to lack ancestral diversity.^38^

## CONCLUSION

Sex-specific differences in risk factors for poor outcomes in HCM mostly relate to heart failure or death. Although sarcomere-negative HCM patients tend to have a milder course, sarcomere-negative women have an increased risk of heart failure later in life, similar to the general female population. Further, women with *MYBPC3* variants have lower risk of heart failure than men with similar variants, the mechanism for which is currently unclear. Additionally, we show sex-specific differences in the performance of the ESC HCM risk score. Further work to explore sex differences in risk of heart failure and death in HCM include collection of female-specific variables such as menarche, pregnancy and menopause, and more detailed comorbidity data. Reporting of sex-disaggregated data are a critical first step in addressing sex disparities and informing sex-specific management strategies.

## PERSPECTIVES

### Competencies in medical knowledge and patient care

- Given men and women have different fundamental underlying biology and exist within societal norms that are heavily gendered, evaluating sex-specific differences in risk factors for poor clinical outcomes is necessary. With a better understanding of this, we can provide tailored insights for clinical practice that incorporate sex-specific risk factors and ultimately benefit both men and women with HCM.

### Translational outlook

- Future research to understand sex-specific risk factors contributing to poor clinical outcomes in HCM is needed, including comprehensive collection of data relating to potential novel risk factors such as age at menarche, pregnancy and menopause. Presentation of sex-disaggregated data is critical in reducing the sex disparity gap.

## Data Availability

All data produced in the present study are available upon reasonable request to the authors and according to ethical approvals

## SOURCES OF FUNDING

The Sarcomeric Human Cardiomyopathy Registry (SHaRe) is supported by unrestricted research grants from Bristol Myers Squibb, Pfizer, inc, and Cytokinetics, including funds to individual sites for database support. No sponsor played a role in the design or conduct of this study nor participated in the article’s preparation, review, or approval. JI is the recipient of a National Health and Medical Research Council (NHMRC) Career Development Fellowship (#1162929). CS is the recipient of a National Health and Medical Research Council (NHMRC) Practitioner Fellowship (#1154992) and a New South Wales Health Cardiovascular Disease Clinical Scientist Grant. This study was partly funded by a New South Wales (NSW) Health Cardiovascular Research Capacity Program early-mid career research grant.

## DISCLOSURES

JI receives research grant support from Bristol Myers Squibb. JWR consultant for Bayer, Merk, Bristol Myers Squibb, Cytokinetics. CA has received honoraria from Astra Zeneca and Amgen. NL is a consultant for Bristol Myers Squibb, Cytokinetics, Pfizer, Tenaya and Sarepta and receives grant support from Pfizer. CYH is a consultant for Bristol Myers Squibb, Cytokinetics, and Viz-AI and receives research support from Bristol Myers Squibb, Cytokinetics, and Pfizer. All remaining authors have nothing to disclose.

## ABBREVIATIONS

HCM: Hypertrophic cardiomyopathy
SHaRe: Sarcomeric Human Cardiomyopathy Registry
AF: Atrial fibrillation
ICD: Implantable cardioverter defibrillator
LV: Left ventricular
P/LP: Pathogenic/Likely pathogenic
VUS: Variant of unknown significance
NYHA: New York Heart Association
LVOTO: Left ventricular outflow tract obstruction
LA: Left atrial

## Notes

### Author Declarations

Ethical approval requiring participant informed consent was granted by Cincinnati Children's Hospital, United States of America (USA); Children's Hospital of Philadelphia, USA; Michigan Medical, USA; Yale Medical, USA; Royal Brompton Hospital, United Kingdom; Erasmus University Medical Center, The Netherlands; Florence Centre for Cardiomyopathies, Italy; and Sydney Local Health District, Royal Prince Alfred Hospital, Australia; InCor, Heart Institute, University of Sao Paulo, Brazil institutional review boards. Ethical approval with a waiver of consent was granted by Stanford School of Medicine, USA; Brigham and Women's Hospital, USA; Boston Children's Hospital, USA; Pennsylvania University Medical Center, USA; and Sydney Local Health District, Royal Prince Alfred Hospital, Australia institutional review boards.

