## Supplementary Material for "Sex-Specific Clinical and Genetic Factors Associated with Adverse Outcomes in Hypertrophic Cardiomyopathy"

**Supplemental Table 1:** Univariable and Multivariable analysis of outcomes in HCM

**Supplemental Table 2:** Characteristics of hypertrophic cardiomyopathy probands at first encounter by sex and sarcomere status

**Supplemental Table 3:** Incident medication use and interventions by sex and sarcomere status

**Supplemental Figure 1:** Sex-disaggregated analysis showing associations of clinical and genetic features with atrial fibrillation

Longitudinal analysis of echocardiography parameters over time by sex

**Supplemental Figure 2:** Longitudinal changes in echocardiographic parameters over time (years) by sex

Page 2

Page 4

Page 5

Page 6

Page 7

Page 8

**SUPPLEMENTAL TABLE 1: Univariate and Multivariate analysis of outcomes in HCM**

| **Covariate** | **Univariate analysis** | | | **Multivariate analysis** | | |
| --- | --- | --- | --- | --- | --- | --- |
|  | **HR** | **95% CI** | **p-value** | **HR** | **95% CI** | **p-value** |
| **Heart failure composite** |  | | | **Events n=353** | | |
| Female sex | 1.77 | 1.56-1.99) | <0.0001 | 1.51 | 1.21-1.88 | 0.0003 |
| Age | 1.08 | 1.07-1.10 | <0.0001 | 1.08 | 1.06-1.11 | <0.0001 |
| Sarcomere positive | 1.02 | 0.87-1.20 | 0.829 | 1.20 | 0.95-1.51 | 0.137 |
| Indexed baseline maximum left ventricular hypertrophy (LVH) | 1.04 | 1.02-1.05 | <0.0001 |  |  |  |
| Indexed left ventricular end-diastolic diameter (LVEDD) | 1.00 | 0.99-1.02 | 0.834 |  |  |  |
| Indexed left ventricular end-systolic diameter (LVESD) | 1.03 | 1.01-1.05 | 0.009 |  |  |  |
| Left ventricular ejection fraction (LVEF) <50% | 2.26 | 1.87-2.73 | <0.0001 | 2.67 | 1.83-3.91 | <0.0001 |
| Indexed left atrial (LA) size | 1.04 | 1.03-1.05 | <0.0001 | 1.04 | 1.02-1.07 | 0.0001 |
| Left ventricular outflow tract obstruction (LVOTO) | 1.90 | 1.65-2.19 | <0.0001 | 1.73 | 1.38-2.17 | <0.0001 |
| Mild-severe mitral regurgitation | 1.28 | 1.12-1.48 | 0.0004 |  |  |  |
| Hypertension | 1.29 | 1.13-1.47 | 0.0002 | 1.14 | 0.90-1.45 | 0.276 |
| Body mass index (BMI) | 1.05 | 1.03-1.06 | <0.0001 | 1.05 | 1.03-1.07 | <0.0001 |
| Atrial fibrillation (AF) | 1.55 | 1.37-1.75 | <0.0001 |  |  |  |
| **Death** |  | | | **Events= 129** | | |
| Female sex | 1.22 | 1.03-1.45 | 0.020 | 0.96 | 0.66-1.41 | 0.843 |
| Age | 0.97 | 0.96-0.97 | <0.0001 | 0.96 | 0.94-0.97 | <0.0001 |
| Sarcomere positive | 1.46 | 1.17-1.83 | 0.001 | 1.26 | 0.85-1.88 | 0.254 |
| Indexed baseline maximum LVH | 1.03 | 1.00-1.06 | 0.026 |  |  |  |
| Indexed LVEDD | 1.02 | 1.01-1.04 | 0.006 |  |  |  |
| Indexed LVESD | 1.03 | 1.02-1.05 | 0.0002 |  |  |  |
| LVEF <50% | 2.80 | 2.22-3.54 | <0.0001 | 2.04 | 1.14-3.65 | 0.0164 |
| Indexed LA size | 1.04 | 1.03-1.06 | <0.0001 | 1.08 | 1.05-1.12 | <0.0001 |
| LVOTO | 1.25 | 1.03-1.53 | 0.028 | 1.89 | 1.27-2.80 | 0.0017 |
| Mild-severe mitral regurgitation | 1.19 | 0.97-1.46 | 0.105 |  |  |  |
| Hypertension | 0.60 | 0.51-0.72 | <0.0001 |  |  |  |
| BMI | 1.01 | 1.00-1.03 | 0.173 |  |  |  |
| **Atrial fibrillation** |  | | | **Events=360** | | |
| Female sex | 0.97 | 0.84-1.10 | 0.6 | 0.74 | 0.59-0.93 | <0.0001 |
| Age | 1.01 | 1.00-1.02 | 0.008 | 1.00 | 0.99-1.02 | 0.608 |
| Sarcomere positive | 1.30 | 1.10-1.54 | 0.002 | 1.48 | 1.17-1.86 | 0.0009 |
| Indexed baseline maximum LVH | 1.03 | 1.01-1.05 | 0.007 |  |  |  |
| Indexed LVEDD | 0.98 | 0.96-1.00 | 0.01 |  |  |  |
| Indexed LVESD | 0.99 | 0.97-1.01 | 0.3 |  |  |  |
| LVEF <50% | 1.40 | 1.09-1.81 | 0.01 |  |  |  |
| Indexed LA size | 1.05 | 1.04-1.06 | <0.0001 | 1.06 | 1.04-1.09 | <0.0001 |
| LVOTO | 1.49 | 1.28-1.73 | <0.0001 | 1.63 | 1.31-2.03 | <0.0001 |
| Mild-severe mitral regurgitation | 1.38 | 1.19-1.61 | <0.0001 |  |  |  |
| Hypertension | 1.15 | 1.00-1.33 | 0.05 | 1.07 | 0.95-1.35 | 0.557 |
| BMI | 1.04 | 1.03-1.05 | <0.0001 |  |  |  |
| **VA composite** |  | | | **Events= 100** | | |
| Female sex | 0.85 | 0.65-1.12 | 0.244 | 0.60 | 0.38-0.94 | 0.027 |
| Age | 0.99 | 0.98-1.01 | 0.7 | 0.99 | 0.97-1.01 | 0.335 |
| Sarcomere positive | 1.61 | 1.16-2.23 | 0.004 | 1.85 | 1.16-2.95 | 0.01 |
| ESC SCD Fhx | 1.59 | 1.08-2.34 | 0.03 | 1.52 | 0.88-2.64 | 0.136 |
| Syncope | 2.14 | 1.63-2.82 | <0.0001 | 1.52 | 0.98-2.36 | 0.063 |
| Indexed baseline maximum LVH | 1.04 | 1.02-1.07 | 0.003 |  |  |  |
| Indexed LVEDD | 1.00 | 0.97-1.02 | 0.6 |  |  |  |
| Indexed LVESD | 1.00 | 0.97-1.03 | 0.966 |  |  |  |
| LVEF <50% | 1.96 | 1.32-2.91 | 0.002 |  |  |  |
| Indexed LA size | 1.02 | 0.99-1.04 | 0.174 | 1.02 | 0.98-1.06 | 0.421 |
| LVOTO | 1.23 | 0.91-1.66 | 0.175 | 1.44 | 0.95-2.18 | 0.088 |
| Mild-severe mitral regurgitation | 1.12 | 0.82-1.49 | 0.508 |  |  |  |
| Hypertension | 0.76 | 0.57-1.02 | 0.0661 |  |  |  |
| BMI | 1.03 | 1.01-1.05 | 0.00351 |  |  |  |

**SUPPLEMENTAL TABLE 2: Characteristics of hypertrophic cardiomyopathy probands at first encounter by sex and sarcomere status**

|  | **Sarcomere-negative** | | **Sarcomere-positive** | |
| --- | --- | --- | --- | --- |
|  | ***Female*** | ***Male*** | ***Female*** | ***Male*** |
|  | 663 | 1245 | 728 | 1047 |
| Age at diagnosis (years) | 54.5 ± 18.4 | 46.9 ± 17.1 | 38.7 ± 18.5 | 35.1 ± 16.4 |
| Follow-up time (years) | 6.5 ± 6.1 | 6.6 ± 6.5 | 8.5 ± 7.7 | 9.1 ± 8.9 |
| Hypertension | 385 (58.1) | 646 (51.9) | 201 (27.6) | 281 (26.6) |
| Body mass index (kg/m^2^) | 28.4 ± 6.7 | 28.8 ± 5.8 | 26.7 ± 6.6 | 26.6 ± 6.7 |
| NYHA III-IV | 135 (24.4) | 99 (9.5) | 101 (16.7) | 87 (10.4) |
| Syncope | 110 (16.7) | 169 (13.9) | 149 (20.7) | 176 (17.3) |
| **Echo characteristics** |  |  |  |  |
| Maximum LV wall thickness, mm | 17.6 ± 5.7 | 18.1 ± 6.0 | 18.9 ± 6.5 | 20.3 ± 6.4 |
| Indexed Max LV wall thickness, mm^a^ | 9.9 ± 3.5 | 8.9 ± 3.7 | 11.0 ± 4.5 | 10.6 ± 4.4 |
| LV end diastolic diameter, mm | 42.0 ± 7.8 | 46.1 ± 6.6 | 41.8 ± 6.5 | 44.6 ± 7.2 |
| Indexed LV end diastolic diameter, mm^a^ | 23.8 ± 5.9 | 22.7 ± 5.5 | 23.9 ± 5.0 | 22.9 ± 6.5 |
| LV end systolic diameter, mm | 25.0 ± 6.7 | 27.8 ± 6.3 | 25.6 ± 6.4 | 27.5 ± 6.9 |
| Indexed end systolic diameter, mm^a^ | 14.0 ± 4.6 | 13.7 ± 4.3 | 14.6 ± 4.2 | 14.1 ± 5.6 |
| LV ejection fraction (%) | 66.3 ± 9.3 | 65.0 ± 8.4 | 64.4 ± 10.0 | 64.0 ± 9.8 |
| LV ejection <50% | 31 (5.3) | 58 (5.3) | 52 (8.7) | 65 (7.3) |
| LV ejection fraction< 35% | 5 (0.9) | 9 (0.8) | 10 (1.7) | 16 (1.9) |
| LV outflow tract obstruction | 305 (58.5) | 399 (44.1) | 183 (32.1) | 212 (25.2) |
| LV outflow tract gradient at rest (mmHg) | 44.1 ± 40.9 | 30.6 ± 32.5 | 27.2 ± 31.5 | 21.0 ± 25.8 |
| Mild-severe mitral regurgitation | 428 (71.5) | 672 (61.7) | 392 (65.2) | 508 (60.2) |
| Left atrial diameter, mm | 42.1 ± 10.2 | 43.3 ± 10.6 | 41.2 ± 10.4 | 43.6 ± 11.4 |
| Indexed left atrial diameter, mm^a^ | 23.9 ± 6.5 | 21.2 ± 5.2 | 23.5 ± 5.8 | 22.3 ± 6.5 |

Data shown as n (%) or mean ± SD.

NYHA indicates New York Heart Association; LV, left ventricular.

^a^Adjusted for body surface area

**SUPPLEMENTAL TABLE 3: Incident medication use and interventions by sex and sarcomere status**

|  | **Sarcomere-negative** | | **Sarcomere-positive** | |
| --- | --- | --- | --- | --- |
|  | ***Female*** | ***Male*** | ***Female*** | ***Male*** |
| **Medications** | 663 | 1245 | 728 | 1047 |
| Beta Blocker | 452 (69.5) | 758 (63.1) | 425 (63.6) | 624 (65.8) |
| Anti-arrhythmic | 70 (13.7) | 119 (12.3) | 108 (19.7) | 150 (20.1) |
| Calcium channel blocker | 175 (26.9) | 249 (20.8) | 140 (21.0) | 167 (17.7) |
| Disopyramide | 72 (11.8) | 84 (7.6) | 56 (9.5) | 57 (6.8) |
| Loop diuretic | 52 (21.0) | 53 (10.3) | 56 (16.3) | 46 (9.3) |
| **Interventions** |  |  |  |  |
| ICD implantation | 118 (17.8) | 266 (21.4) | 293 (40.3) | 412 (39.4) |
| Alcohol septal ablation | 53 (8.0) | 46 (3.7) | 37 (5.1) | 35 (3.4) |
| Septal myectomy | 157 (23.7) | 233 (18.7) | 149 (20.5) | 152 (14.5) |

Data shown as n (%). ICD indicates implantable cardioverter defibrillator

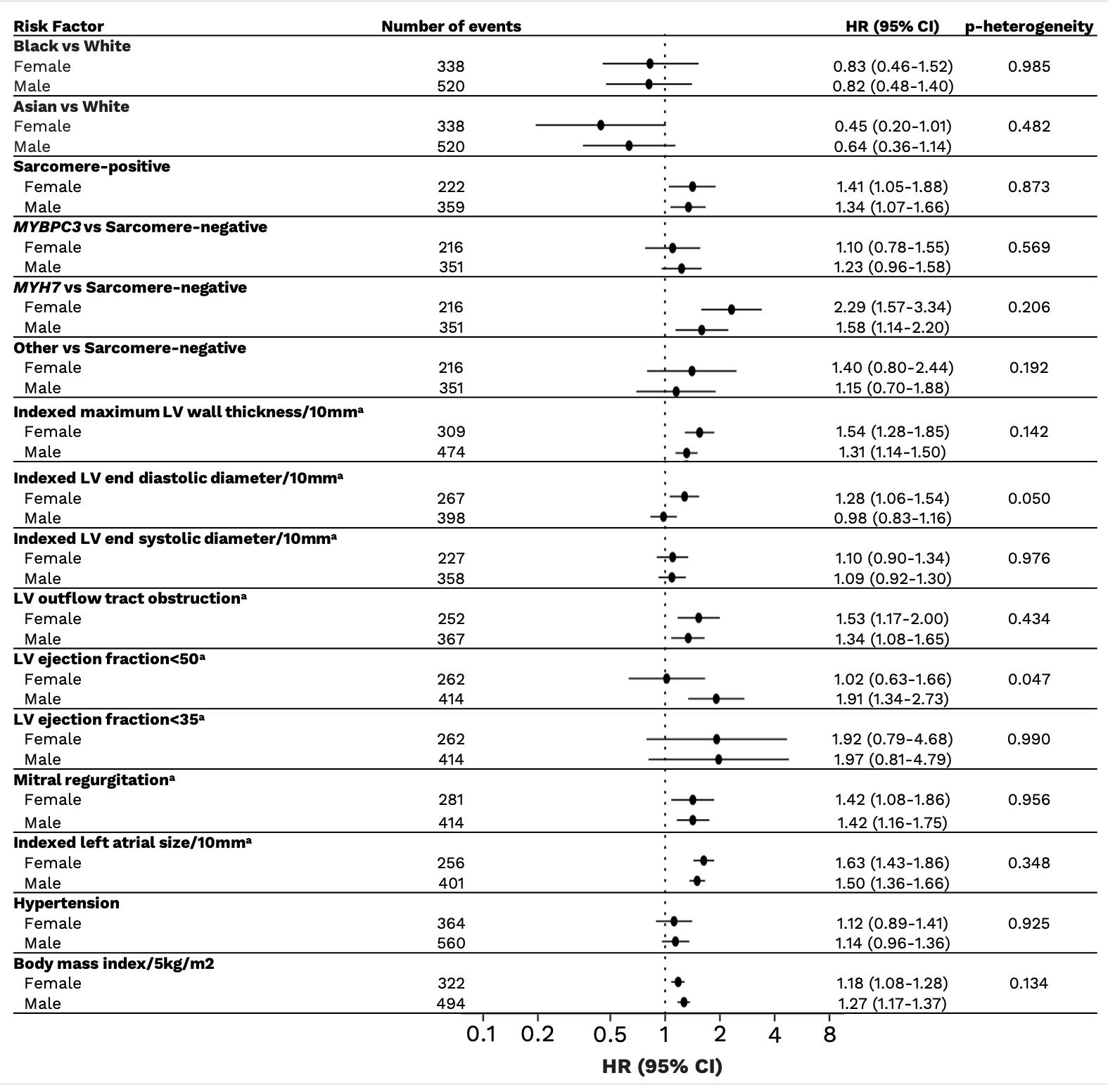
**Supplemental Figure 1: Sex-disaggregated analysis showing associations of clinical and genetic features with atrial fibrillation**

^a^Echocardiography measurements adjusted for body surface area

Abbreviations: CI; confidence interval; HR; hazard ratio; LV, left ventricular.

**Longitudinal analysis of echocardiography parameters over time by sex**

Longitudinal analysis was conducted on echocardiography parameters, indexed maximum left ventricular (LV) wall thickness and left atrial (LA) size, to investigate whether disease progression over time differed between sexes. We used linear mixed models (R package lme4 v1.1.29) to compare women and men over time after rank-normalizing the parameters of interest. The echocardiography variable of interest was used as the outcome variable, and we tested for interactions between sex and age (measured in years). Additionally, all models included a random effect to account for repeated observations from the same individuals over time and parameters site, sarcomere status, obstruction and body surface area were included as fixed effects. We found no significant interaction for maximum LV wall thickness (anova comparison, p>0.05; Supplemental Figure 2A), but we did find a significant interaction for LA size (anova comparison, p=0.018). Further analysis of this result by age categories found a significant association for 20-29 years and 60-69 years (anova comparison, p=0.010 and p=0.0003, respectively; Supplemental Figure 2B). No significant interaction existed for other age groups. While we have shown a significant association, we are mindful of the weakness in using this dataset given the intra-test variability within individuals, which may not represent their underlying HCM. Better-controlled experiments would be necessary to confirm these results.

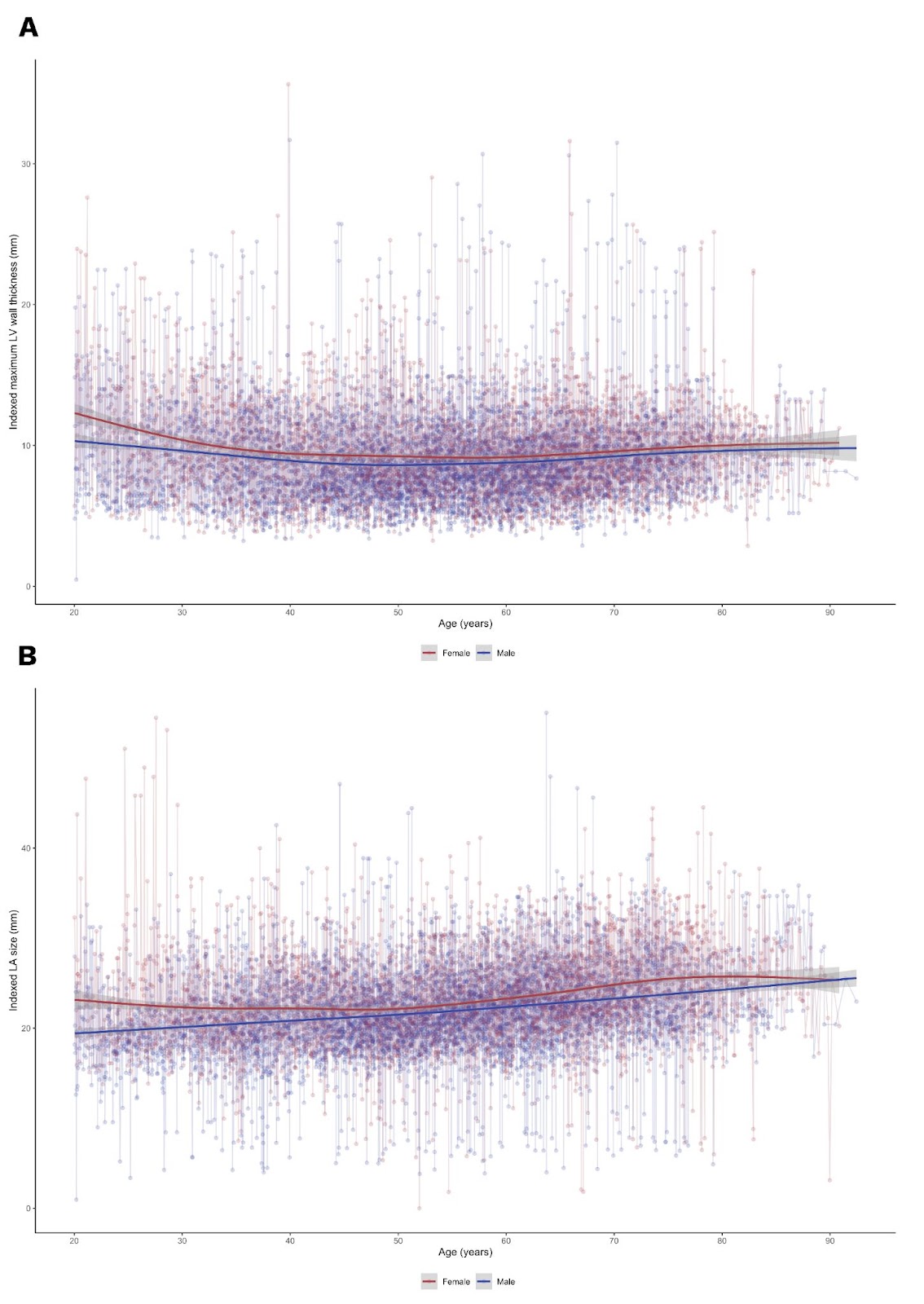

**SUPPLEMENTAL FIGURE 2: Longitudinal changes in echocardiographic parameters over time (years) by sex. A: Indexed maximum left ventricular (LV) wall thickness B: Indexed left atrial (LA) size**
